# Medial prefrontal transcranial alternating current stimulation for apathy in Huntington’s disease

**DOI:** 10.1101/2022.08.29.22279310

**Authors:** Marie-Claire Davis, Aron T. Hill, Paul B. Fitzgerald, Neil W. Bailey, Caley Sullivan, Julie C. Stout, Kate E. Hoy

## Abstract

We investigated the effects of transcranial alternating current stimulation (tACS) targeted to the bilateral medial prefrontal cortex (mPFC) and administered at either delta or alpha frequencies, on brain activity and apathy in people with Huntington’s disease (HD) (n = 17). Neurotypical controls (n = 20) were also recruited for comparison. All participants underwent three 20-minute sessions of tACS; one session at alpha frequency (Individualised Alpha Frequency (IAF), or 10Hz when an IAF was not detected); one session at delta frequency (2Hz); and a session of sham tACS. Participants completed the Monetary Incentive Delay (MID) task with simultaneous recording of EEG immediately before and after each tACS condition. The MID task presents participants with cues signalling potential monetary gains or losses that increase activity in key regions of the cortico-basal ganglia-thalamocortical networks, with dysfunction of the latter network being implicated in the pathophysiology of apathy. We used the P300 and Contingent Negative Variation (CNV) event-related potentials elicited during the MID task as markers of mPFC engagement. HD participants’ CNV amplitude significantly increased in response to alpha-tACS, but not delta-tACS or sham. Neurotypical controls’ P300 and CNV were not modulated by any of the tACS conditions, but they did demonstrate a significant decrease in post-target response times following alpha-tACS. We present this as preliminary evidence of the ability of alpha-tACS to modulate brain activity associated with apathy in HD.

## 1.1 Introduction

Huntington’s disease (HD) is a neurodegenerative disease caused by expansion of the trinucleotide repeat (cytosine adenine guanine; CAG) in the *huntingtin (HTT*) gene on chromosome 4 (Pender and Koroshetz 2011, Podvin et al. 2019). Apathy is a highly prevalent and functionally debilitating neuropsychiatric symptom in HD, for which there are currently no clinically effective treatments (Fritz et al. 2018, Jacobs, Hart, and Roos 2018, Martinez-Horta et al. 2016, Paoli et al. 2017). Degeneration and dysfunction within the cortico-basal ganglia-thalamocortical (CBGT) networks contributes to the motor, cognitive and psychiatric symptoms in HD, including apathy (Papoutsi et al. 2014, Ross et al. 2014, Ross et al. 2017).

Apathy in conditions affecting the CBGT networks (e.g., Parkinson’s disease and schizophrenia) is associated with abnormal oscillatory activity within the delta and alpha frequency bands (Hatz et al. 2017, Bortolon et al. 2017, Knyazev 2012, Schutter and Knyazev 2012, Le Heron, Apps, and Husain 2017, Lanctôt et al. 2017). Similarly, lower alpha power and higher delta power in people with HD, relative to neurotypical controls, has also been a consistent finding (Delussi et al. 2020, Hunter et al. 2010, Odish et al. 2018, Painold et al. 2010, Piano, Imperatori, et al. 2017, Piano, Mazzucchi, et al. 2017, Ponomareva et al. 2014). Neuroanatomically, dysfunction in bilateral anterior cingulate and medial prefrontal cortex (mPFC) is common across conditions causing apathy, including HD (Le Heron, Apps, and Husain 2017, Lanctôt et al. 2017). These pathological differences suggest potential therapeutic targets for the treatment of HD-related apathy using non-invasive brain stimulation (NIBS).

Initial investigations into NIBS for the treatment of apathy in other neurological conditions have demonstrated some promising behavioural results (Padala et al. 2018, Sasaki et al. 2017, although see Suemoto et al. 2014); however, no studies have been conducted in HD and none have directly investigated biological target engagement, which is considered best practice for the development of brain stimulation treatments (Insel 2015).

Transcranial alternating current stimulation (tACS) is a NIBS technique that can target pathophysiologically relevant oscillatory frequencies within specific regions of the cortex (Thut, Schyns, and Gross 2011, Tavakoli and Yun 2017, Onoda et al. 2017). The immediate (‘online’) effects of tACS are considered to reflect exogenous entrainment of endogenous oscillatory activity and stochastic resonance (Fertonani and Miniussi 2017, Thut, Schyns, and Gross 2011, Huang, Lu, et al. 2017, Liu et al. 2018), while post-stimulation (‘offline’) effects are considered secondary to spike timing dependent plasticity (Herrmann et al. 2013, Vossen, Gross, and Thut 2015, Vogeti, Boetzel, and Herrmann 2022). The post-stimulation effects on behaviour following a single 20-minute session of tACS may last as long as 70 minutes (Kasten, Dowsett, and Herrmann 2016), with daily sessions over a period of one to several weeks able to engender clinically meaningful outcomes (Elyamany et al. 2021). tACS also benefits from being economical, portable (e.g., can fit in a small case), and easy to use, enabling home-based use by appropriately trained participants for both experimental and clinical research purposes (Charvet et al. 2018, Charvet et al. 2015, Palm et al. 2018). For the current study, we chose to administer tACS at either delta or alpha frequencies, based on the aforementioned evidence of abnormal delta and alpha oscillatory activity in HD and other conditions affecting the CBGT networks in which apathy is prevalent.

To assess the capacity of tACS to modify activity in the mPFC, we investigated its effects on event-related potentials elicited using the EEG version of the Monetary Incentive Delay (MID) task (Knutson et al. 2000, Broyd et al. 2012). We chose the MID task because of its conceptual and neuroanatomical relevance to apathy. The MID task presents participants with motivationally salient cues signifying that they can gain or lose money depending on the speed of their subsequent behavioural responses (i.e., pressing a button in response to the presentation of a target cue). Motivationally salient cue processing is one of several neurocognitive subprocesses thought to underpin motivated, and reciprocally, amotivated or apathetic behaviour (Chong 2018, Husain and Roiser 2018, Kringelbach and Berridge 2016). That is, perceiving cues in our environment as motivationally salient or otherwise, influences how we direct our cognitive, emotional, and physical resources (Bromberg-Martin, Matsumoto, and Hikosaka 2010, Olney et al. 2018). The MID task’s neuroanatomical relevance has been demonstrated by extensive implementation of the task concurrent with fMRI, where motivationally salient cues have been shown to elicit significantly greater activation in CBGT networks than ‘neutral’ cues (Dugré et al. 2018, Wilson et al. 2018, Oldham et al. 2018). Participants with premanifest HD and Parkinson’s disease have also been found to demonstrate abnormal activation patterns on the fMRI version of the MID task (Enzi et al. 2012, du Plessis et al. 2018). Research using the EEG version of the MID task has shown that event-related potentials (ERPs) elicited by cue presentation, specifically the P300 and Contingent Negative Variation (CNV), also vary according to the motivational salience of the cue (Hill et al. 2018, Novak and Foti 2015, Pfabigan et al. 2014, Zhang et al. 2017). As such, we used the P300 and CNV potentials elicited by the EEG version of the MID task as neurophysiological markers of apathy.

In summary, we conducted a within-subjects comparison of three tACS conditions (alpha-tACS; delta-tACS; sham-tACS) across three separate sessions in people with HD and neurotypical controls. We hypothesised that a single 20-minute session of tACS targeting frequencies in the alpha or delta frequency bands would, relative to sham, modulate neural activity, particularly within the mPFC. We hypothesised that this would increase HD participants’ P300 and CNV amplitude following motivationally salient cues elicited during the MID task. We predicted similar effects on P300 and CNV amplitude in neurotypical controls following either alpha or delta tACS.

## 2.1 Materials and Method

### 2.2 Participants

We recruited 22 participants with HD and 20 neurotypical controls aged between 18 and 75 years (Australian New Zealand Clinical Trials Registry (ANZCTR): 12619000870156). Data collection took place between February 2019 and August 2021. This study received ethics approval from the Alfred Health, Calvary Health Care Bethlehem, and Monash University Human Research Ethics Committees, and all participants gave their written informed consent. This paper presents the results of tACS effects on task-related EEG and behavioural data. Please note that we have previously analysed the baseline differences between our participants with HD (n = 22) and neurotypical controls (n = 20) to explore the relationship between neurophysiological markers and clinical measures of apathy and cognition (Davis, Hill, Fitzgerald, Bailey, et al. 2022, Davis, Hill, Fitzgerald, Stout, et al. 2022).

The participants with HD were all confirmed to have the HD CAG expansion, and in the late premanifest (n = 11) or early manifest stage 1 (n = 11) of their disease, with a disease burden score (DBS) of ≥280 (DBS = [CAG repeats - 35.5] x current age) and all had a Total Functional Capacity score (TFC) of ≥ 9 (maximum score of 13) (Ghosh and Tabrizi 2018, Langbehn et al. 2004, Kieburtz 1996, Penney et al. 1997). The TFC is a clinician rating of how the person with HD is functioning across the domains of occupation, finances, domestic chores, activities of daily living, and care level (Beglinger et al. 2010, Shoulson and Fahn 1979). Our choice to recruit people in the late premanifest and early manifest stages of the disease continuum reflected the prevalence and functional impact of apathy and mild cognitive impairment during these stages, when the person with HD is continuing to participate in home, work, and social activities (Fritz et al. 2018, Jacobs, Hart, and Roos 2018, Van Der Zwaan et al. 2021).

Study exclusion criteria were as follows: Use of anticonvulsant medications or benzodiazepines; commencement or change in dose of other psychotropic medications (i.e., anti-depressants, anti-psychotics) during the four weeks prior to, and/or during participation; choreiform movements preventing EEG data collection; a current substance use or alcohol use disorder or episode of psychiatric illness, as assessed and defined by the Mini International Neuropsychiatric Interview (MINI) 7.0.2 (Sheehan et al. 1997); or a previous head injury or traumatic brain injury defined by a loss of consciousness >30 minutes or requiring a hospital admission.

There were significant differences between the late premanifest (n = 11) and early manifest (n = 11) groups with regards to total motor symptoms, TFC, processing speed, and average age, but they did not differ significantly in average DBS scores (Manifest DBS M(SD) = 370.7(30.3), Range = 283.5-495.0; Late premanifest DBS M(SD) = 351.0(60.3), Range = 286.0-484.5, *t20 = -0*.*77, p = 0*.*45*). There were also no significant differences between the groups on the measures of general cognition, apathy, anxiety, depression, years of education, or whether they used psychotropic medications (refer to Table s1 in the supplementary materials for details of all comparisons). The highest total motor score of 49 for the combined HD group was also relatively mild (given the maximum score is 131). These similarities supported our decision to use a combined sample and aligns with the increasing recognition of non-motor onset and cognitive-neuropsychiatric phenotypes in HD (Kim et al. 2014, Mehrabi et al. 2016, Ross et al. 2019, Paulsen et al. 2017). Fourteen participants with HD were taking stable doses of psychotropic medications comprising SSRIs (n = 8), risperidone (n=7), mirtazapine (n = 3), SNRIs (n = 2), and tetrabenazine (n = 1). One of the control participants was on a low dose tricyclic antidepressant (amitriptyline) as a migraine prophylactic. Refer to Table 1 for all demographic and clinical information.

**Table 1.**
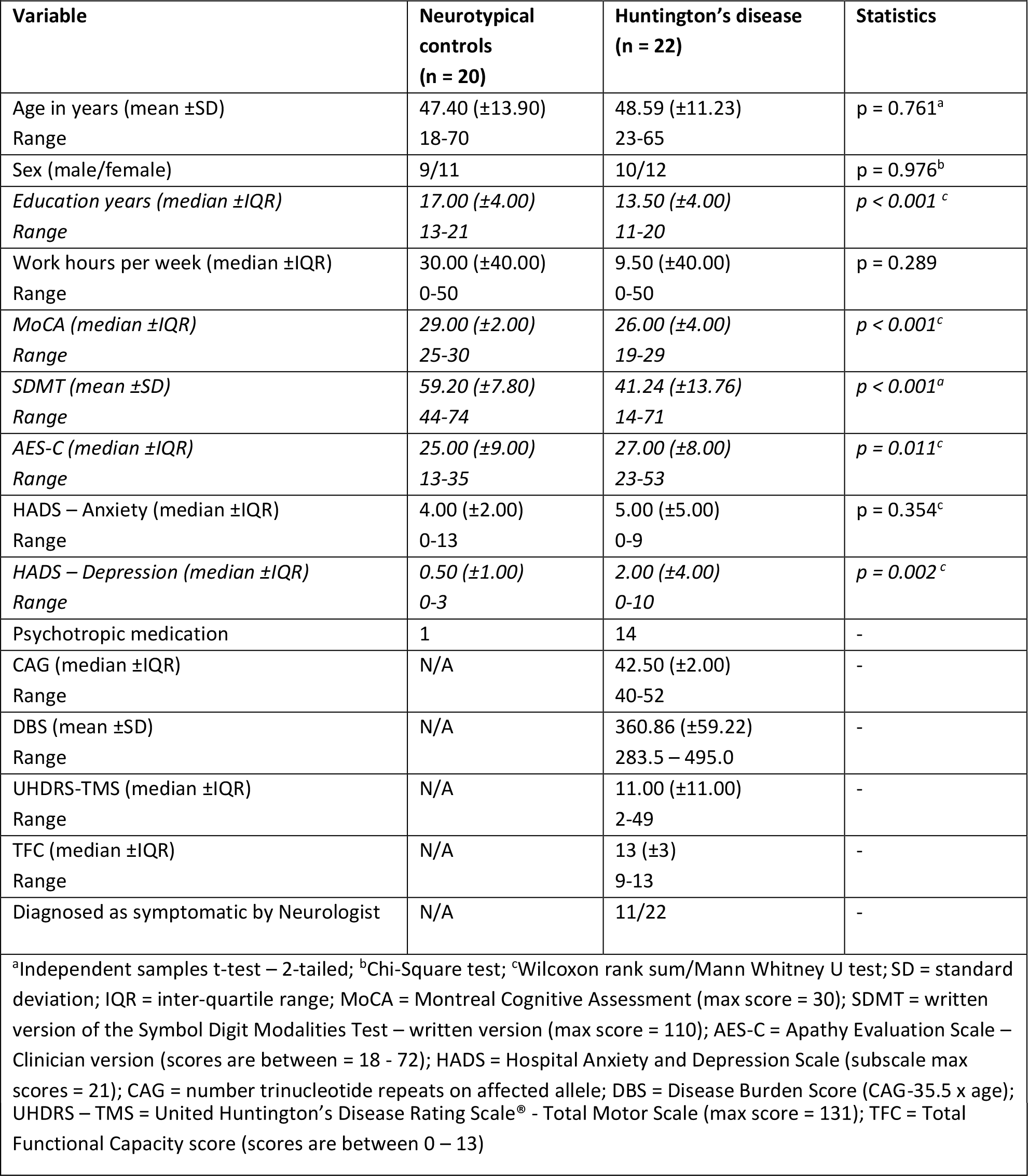
Characteristics of participant groups.

### 2.3 Sample characterisation measures

The sample characterisation measures were as follows: The Unified Huntington’s Disease Rating Scale (UHDRS^®^) total motor score (clinician rated; Kieburtz 1996); the Montreal Cognitive Assessment (MoCA), which is a rater-administered brief cognitive screen (Nasreddine et al. 2005); the Hospital Anxiety and Depression Scale (HADS), a self-report measure of symptoms of anxiety and depression, with scores of ≤7 on both subscales considered to be within the ‘normal’ range (Zigmond and Snaith 1983, Stern 2014); the Apathy Evaluation Scale – clinician version (AES-C), which is a semi-structured clinical interview to assess the changes in overt behaviour, emotional responsiveness and goal-directed thought content associated with apathy (Marin, Biedrzycki, and Firinciogullari 1991); and the written version of the Symbol Digit Modalities Test (SMDT), which is a rater-administered measure of processing speed well-validated for use in HD research (Smith 1982, Stout et al. 2014). All measures, including the UHDRS^®^ total motor scale, were administered by MCD, an experienced clinical neuropsychologist with certification in administration of the UHDRS^®^ total motor score through the Enroll-HD clinical training portal (https://hdtraining.enroll-hd.org/). COVID-19 social distancing restrictions and lockdowns in Melbourne Victoria impacted data collection for a small number of the participants with HD. Specifically, the UHDRS^®^ total motor score and MoCA data were not collected for three HD participants, and the SDMT and AES-C data are missing for one HD participant. With regards to the tACS and EEG data, one HD participant was unable to attend their alpha-tACS session, one HD participant was unable to attend their sham-tACS session, and three HD participants were unable to attend the session in which they had been allocated to complete delta-tACS. Task-related EEG data (but not behavioural data) was also missing for another HD participant due to a technological failure during their sham-tACS session. The data for these participants was therefore excluded from applicable analyses (i.e., comparisons of all three tACS conditions).

### 2.4 Monetary Incentive Delay task

Participants completed the EEG version of the MID task before and after tACS (i.e., six times in total for the three stimulation conditions [alpha-tACS, delta-tACS, sham-tACS]). We used a version of the task similar in timing parameters and stimuli to previous studies (Novak and Foti 2015, Pfabigan et al. 2014, Zhang et al. 2017). The task was administered via Inquisit Lab version 4 software (Millisecond 2015). Participants were presented with gain, loss and neutral cues signalling that they could expect to gain money, lose money or neither outcome according to the speed of their response to a target stimulus presented a few seconds after each cue.

Specifically, gain cues signalled to the participant that they could win one dollar (+AU$1) if they pressed the enter key quickly enough after seeing a white square (target). If the participant was too slow, they did not win money. Loss cues signalled to the participant that they would *not* lose one dollar if they pressed the enter key quickly enough after seeing a white square. If the participant was too slow to respond to the white square, however, then they *would* lose one dollar (-AU$1). Neutral cues signalled to the participant that they were expected to respond as quickly as possible when they saw a white square but would neither gain nor lose any money during that trial. Feedback was provided after each response (refer to Figure 1), and participants’ cumulative winnings were updated at the base of the screen. The task was structured so that participants completed 10 simple reaction time trials to establish their average baseline reaction time, followed by 12 practice trials (four of each gain, loss, and neutral cue type), and then two blocks of 75 test trials (i.e., 150 test trials) comprising 50 gain cues, 50 loss cues, and 50 neutral cues, presented in random order.

**Figure 1.**
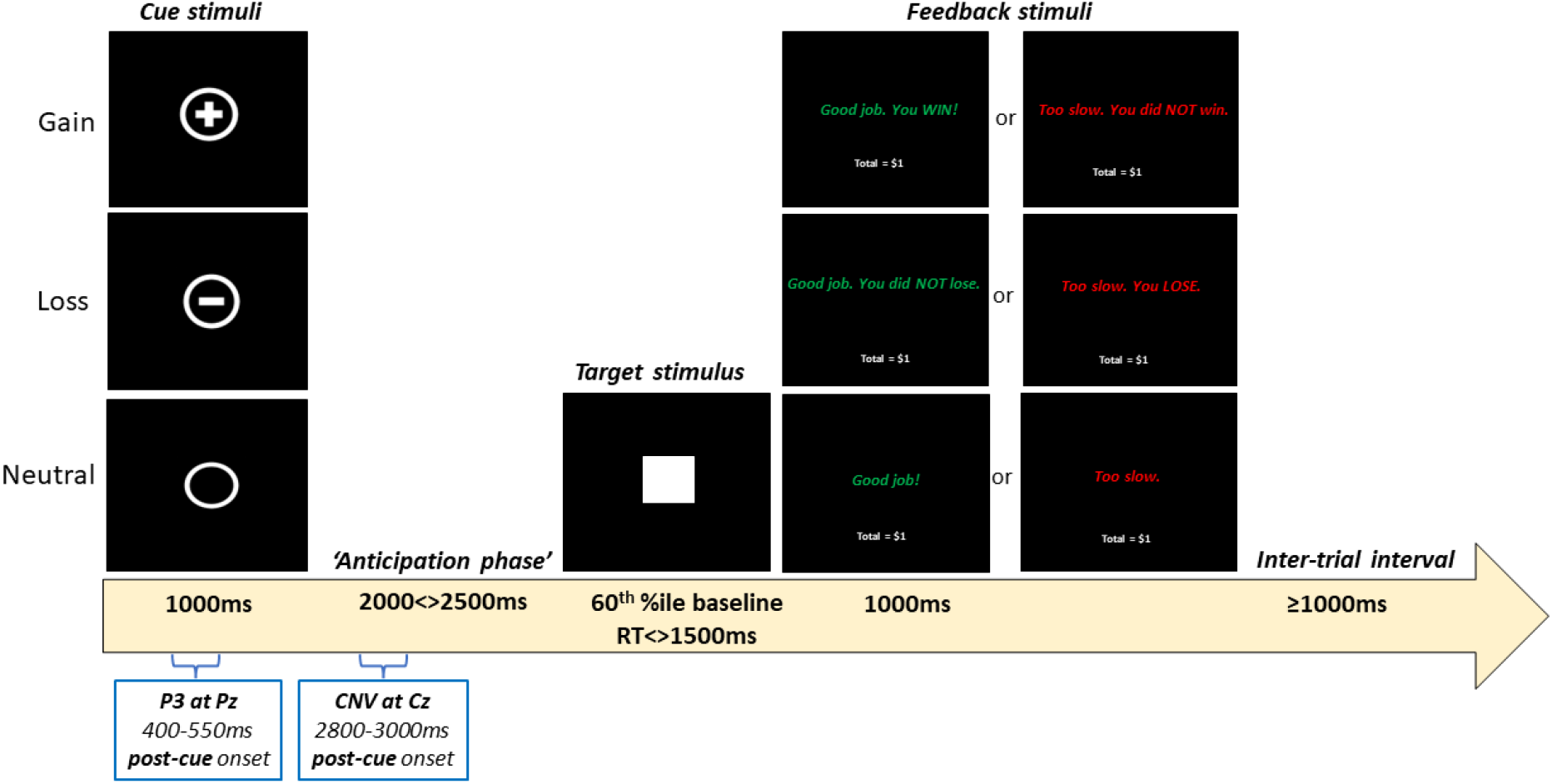
Stimuli and timing parameters for each 7000ms trial of the EEG version of the MID task.

Each trial was 7000ms in duration and began with the cue presented for 1000ms, followed by an “anticipation phase” that varied randomly between 2000-2500ms (in 100ms increments) (refer to Figure 1). The target (white square) was initially presented for a duration corresponding to the 60th percentile of the participant’s average baseline reaction time. The target disappeared as soon as the participant responded. Target response times were recorded for subsequent analysis, with response times below 50ms considered invalid and removed prior to statistical analysis (Pfabigan et al. 2014). To ensure each participant obtained a comparable correct response rate of approximately 66% (Knutson et al. 2000), and to minimize floor and ceiling effects in participants’ response time data, the duration of target presentation increased by 30ms after 2 incorrect (i.e., too slow) responses, or decreased by 30ms after 3 correct (i.e., fast enough) responses using an adaptive algorithm (Hahn et al. 2009). Participants’ individualized target duration could increase to a maximum of 1500ms to accommodate slowed processing speed in participants with HD. Feedback was presented for 1000ms. The intertrial interval was at least 1000ms. Overall task duration was ∼22 minutes. Participants were reimbursed a baseline sum of AU$30 but could earn up to an additional $30 per session, based on their winnings during the MID task, as per the standard administration guidelines to ensure task validity (Knutson et al. 2000).

The P300 was measured as the average amplitude from 400-550ms post-cue at electrode Pz, while the CNV was measured as the average amplitude from 2800-3000ms post-cue at electrode Cz. The P300 is a positive EEG voltage deflection that occurs approximately 300ms after stimulus presentation and detected in frontoparietal midline electrodes (Polich 2012, Huang, Chen, and Zhang 2015). It is functionally associated with increased attentional focus to support memory storage of task-relevant information (Polich 2012, Huang, Chen, and Zhang 2015). The CNV is a negative EEG voltage potential typically measured along midline electrodes and is elicited when a warning stimulus (e.g., a cue) signals the need to respond quickly to a target and is thought to reflect the effect of motivation or effort on motor preparation (Glazer et al. 2018, Zhang and Zheng 2022, de Tommaso et al. 2020).

We chose to use single electrodes to measure each ERP (i.e., Pz for P300 and Cz for CNV) as these were the only electrodes common to previous studies using similar versions of the MID (Broyd et al. 2012, Novak and Foti 2015, Pfabigan et al. 2014, Zhang et al. 2017). Our choice of temporal measurements replicated those of Zhang and colleagues, who used similar cue stimuli and stimulus presentation and anticipation intervals (i.e., cues presented for 1000ms and 2000<>2500ms for the anticipation phase) (Zhang et al. 2017). The period 2800-3000ms post-cue for the CNV was also the latest interval we could measure without potentially impinging on target presentation.

### 2.5 Transcranial alternating current stimulation

Participants underwent all three tACS conditions and were blind to stimulation condition and order. Session order was also randomised. Each session was of approximately three hours’ duration and sessions were completed at least 72 hours apart to ensure no carry-over effects of stimulation. All three sessions were completed within a three-month period to minimise the potential confound of neurological progression among the participants with HD. Stimulation was administered via the Starstim^®^ wireless hybrid EEG/transcranial current stimulation 8-channel system (Neuroelectrics, Barcelona, Spain). At the end of each session, participants were asked to report whether they believed that they had received ‘real’ or ‘sham’ tACS.

#### 2.5.1 tACS conditions

##### 2.5.2 Alpha-tACS

Alpha-tACS was administered at each participant’s peak individualized alpha frequency (IAF) based on evidence that tACS at IAF can increase alpha power more effectively than when administered at a generic frequency, such as 10Hz (Negahbani et al. 2018, Vogeti, Boetzel, and Herrmann 2022). Each participant’s peak IAF was calculated in MATLAB using five minutes of resting eyes open EEG data collected at the start of the session in which they were scheduled to receive alpha-tACS (refer to supplementary materials for IAF detection procedure). During these five minutes of EEG data collection, participants viewed a portion of the same underwater footage they were exposed to while receiving tACS. When an IAF was unable to be found for a participant, they received stimulation at 10Hz.

##### 2.5.3 Delta-tACS

Delta-tACS was administered at 2Hz for all participants. Research using tACS targeting the delta frequency band is limited (Klink et al. 2020), therefore its inclusion in the current study was exploratory. While we aimed to modulate delta oscillatory power, we did not make directional hypotheses regarding the effects of delta-tACS. Our choice of 2Hz was based on the available evidence from experimental human and animal studies (Wischnewski and Schutter 2017, Parker et al. 2017, Kim et al. 2017).

##### 2.5.4 Sham-tACS

Sham-tACS was administered at 10Hz and comprised of 30 seconds of “ramping up” to peak intensity followed immediately by 30 seconds of “ramping down” to zero intensity stimulation.

### 2.6 tACS intensity and duration

Simulation intensity for all conditions was 2mA (peak-to-peak), centred around zero (i.e., with no DC offset). Stimulation duration was limited to 20 minutes per session (including 30 seconds of “ramping up” to 2mA at commencement, and 30 seconds “ramping down” at 19 minutes 30 seconds). Participants were asked to relax, keep their eyes open and focus on a silent video of underwater coral and tropical fish (Undersea Productions, Queensland, Australia) to reduce the likelihood of drowsiness and boredom over this 20-minute period.

### 2.7 tACS montage

The montage targeted bilateral mPFC using circular rubber electrodes affixed to the scalp with conductive paste (Ten20 conductive paste; Weaver and Co., Colorado, USA; contact area 12.56cm2) at the 10-20 system locations of Afz (-2mA), C5 (1mA), and C6 (1mA). This montage was chosen based on computational electric field modelling, which indicated good coverage across brain regions corresponding approximately to the bilateral medial prefrontal cortices. Figure 2 shows the computer modelling output using the open-source ROAST package (Huang, Datta, et al. 2017) that supported the choice of montage and stimulation intensity.

**Figure 2.**
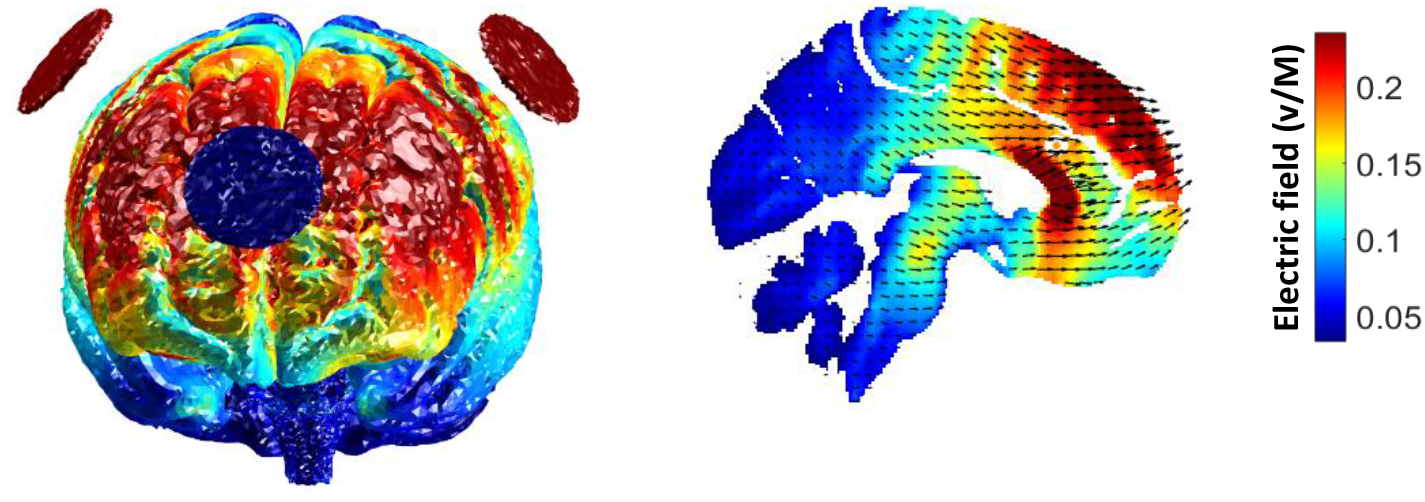
Computer modelling of -2mA at Afz, 1mA at C5 and 1mA at C6.

### 2.8 EEG recording and data preprocessing

EEG data were collected during rest before alpha-tACS to determine IAF and for an analysis of resting EEG (reported separately). EEG data were also collected while participants completed the MID task before and after tACS administration. We used a 45-channel montage (EasyCap, Heersching, Germany) via the Neuroscan EEG system with Ag/AgCl electrodes connected to a SynAmps amplifier (Compumedics, Melbourne, Australia). Task presentation was synchronised with the EEG recording via the Inquisit 4 software, with EEG markers coinciding with the onset of each motivational cue presentation. The electrodes used were: AF3, AF4, F7, F5, F3, F1, Fz, F2, F4, F6, F8, FC5, FC3, FC1, FCz, FC2, FC4, FC6, T7, C3, C1, Cz, C2, C4, T8, CP5, CP3, CP1, CPz, CP2, CP4, CP6, P7, P5, P3, P1, PZ, P2, P4, P6, P8, PO3, POz, PO4, O1, Oz, O2. Recording was completed at a sampling rate of 1KHz, with impedances kept below 5kΩ, and the signal online referenced to CPz, with the ground at FCz. The EEG data were pre-processed offline using RELAX, a fully automated EEG pre-processing pipeline (Bailey, Biabani, et al. 2022, Bailey, Hill, et al. 2022) implemented in MATLAB (MathWorks 2019) and incorporating the EEGLAB toolbox (Delorme and Makeig 2004). Firstly, a notch filter (47<>52Hz) was applied to remove line noise, along with a fourth order Butterworth bandpass filter (0.25<>80Hz) with zero phase (Rogasch et al. 2017). The RELAX pipeline consisted of an initial extreme outlying channel and period rejection step. The initial bad electrode rejection was performed by the PREP pipeline (Bigdely-Shamlo et al. 2015). This was followed by a multi-channel Wiener filter algorithm which provided an initial reduction of eye movement, muscle activity and drift artifacts (Somers, Francart, and Bertrand 2018). Artifacts informing the Wiener filter algorithm were automatically identified using muscle artifact detection methods described by Fitzgibbon and colleagues (2016), then blink, eye movement and drift were identified using amplitude thresholds for correction by the Multiple Wiener filter (Bailey, Biabani, et al. 2022). Remaining artifacts identified via independent component analysis (ICA) using the automated ICLabel classifier (Pion-Tonachini, Kreutz-Delgado, and Makeig 2019) were then removed using wavelet enhanced independent component analysis (wICA). The data were re-referenced to the average of all electrodes and segmented into non-overlapping 6.5-second epochs (-1.5 to 5.0 seconds around cue onset markers). Given that we were investigating ERP responses elicited by cues, all epochs were included for analysis, regardless of whether the participant’s response was ‘correct’ (i.e., they responded quickly enough on presentation of the white square). The average number of epochs for each cue type across the three sessions remaining for analysis were as follows for neurotypical controls: Gain M(SD) = 42.13 (3.97); Loss M(SD) = 42.67 (3.73); Neutral M(SD) = 41.93 (3.83). The remaining epochs for participants with HD: Gain M(SD) = 42.08 (4.03); Loss M(SD) = 42.16 (4.15); and Neutral M(SD) = 41.82 (3.96). All epochs were baseline normalised by subtracting the average amplitude from -500ms to -200ms prior to cue onset from the active period data. Using FieldTrip (Oostenveld et al. 2011), each participants’ epochs from before and after each tACS condition were averaged according to cue type and combined to form grand averages for the HD and neurotypical control groups.

### 2.9 Statistical analyses

Within-group differences in ERP amplitude and response time associated with each cue type (gain, loss and neutral) following each tACS condition were assessed using repeated measures analysis of variance (ANOVA). All pairwise comparisons were Bonferroni corrected. Given analyses were based on *a priori* hypotheses and investigated within a small sample, we did not correct for experiment-wise multiple comparisons. Extreme outliers (i.e., a Z score >±3.0) detected on outcome variables were winsorized (i.e., 90th percentile method) prior to further analysis. Analyses using winsorized data were replicated using the raw dataset and are presented in the supplementary materials for comparison. All statistical analyses were carried out using IBM SPSS version 28.0 (IBM Corp 2021).

## 3.1 Results

### 3.2 Group differences in clinical measures

As depicted in Table 1, the HD participants’ scores on the MoCA and SDMT were significantly lower than that of the neurotypical control group, and the control group had a significantly higher number of years of education than the HD group. The HD group also had scores in the moderate range on the measures of apathy and depression symptoms. Two participants with HD scored 8 and 10 respectively on the HADS depression subscale, putting them in the ‘mild’ (8-10) range for depression. Similarly, three neurotypical controls scored 8, 11 and 13 and one participant with HD scored 9 on the anxiety subscale, putting these participants in the ‘mild’ (8-10) and ‘moderate’ (11-14) ranges for anxiety on this screening measure (Stern 2014). Note however, that none of the participants from either group met criteria for a current psychiatric illness as assessed and defined by MINI.

### 3.3 Detection of Individualised Alpha Frequency

An IAF was only able to be detected in 10 of the 21 HD participants who participated in the alpha-tACS condition (Mean(SD) IAF = 9.79(1.64)Hz; Range = 7.8-12.8 Hz). By contrast, an IAF was detectable for 16 of the 20 control participants (Mean(SD) IAF = 9.99(1.34)Hz; Range = 8.3-12.3 Hz). This group difference was statistically significant (*X2 = 4*.*63, p = 0*.*03*).

We compared baseline characteristics of those HD participants for whom we were able to detect an IAF to those for whom we were not able to detect an IAF. The groups did not differ on the measures of cognition, apathy, symptoms of anxiety or depression, use of psychotropic medications, or motor symptoms (refer to Table s2). These IAF subgroups did, however, differ significantly with regards to their average DBS; the group of HD participants for whom we found an IAF having a significantly lower DBS (Mean DBS IAF group = 334.45; Non-IAF group = 391.41, *t19 = 2*.*51, p = 0*.*02*).

### 3.4 Blinding and side effects

The proportion of participants endorsing that they had received ‘real’ tACS did not significantly differ across the three tACS conditions (alpha = 78%; delta = 69%; sham = 80%; *Cochrane’s Q = 0*.*90, p = 0*.*64*), indicating that blinding of participants was successful. All stimulation sessions were well-tolerated, with participants reporting only mild side effects (e.g., brief itching tingling sensations and phosphene perception) that resolved on cessation of stimulation.

### 3.5 Percentage of correct responses on the MID

We conducted a mixed between-within subjects’ ANOVA comparing percentage of correct responses before and after each tACS condition across the two participant groups to confirm the efficacy of the algorithm manipulating target presentation times. There were no significant main effects or interactions, therefore reinforcement rates were similar for participants in both groups across the six repetitions of the MID task (refer to Tables s3 and s4).

### 3.6 Effect of tACS on CNV amplitude

The effect of tACS on CNV amplitude at electrode Cz following each cue type was assessed using separate within-subjects’ ANOVAs for each group (*tACS:* alpha vs delta vs sham x *cue type:* gain vs loss vs neutral x *time:* pre vs post).

#### 3.6.1 Participants with HD

The ANOVA for the HD group (n = 17) revealed a significant main effect for cue type *(F2*,*15 = 3*.*89, p = 0*.*04, partial eta2 = 0*.*34*), and a significant interaction between tACS and time (F2,15 = 8.61, *p = 0*.*003, partial eta2* = *0*.*53*).

Collapsed across tACS condition and time, pairwise comparisons found that HD participants’ CNV amplitude in response to gain cues was significantly greater than their CNV amplitude in response to loss cues (gain vs loss mean difference = -0.19, *p = 0*.*04*). The other comparisons of CNV amplitude by cue type were non-significant (gain vs neutral mean difference = -0.19, *p = 0*.*21*; loss vs neutral = -0.004, *p = 1*.*00*). This suggests that HD participants’ CNV amplitude was greater in response to gain cues regardless of tACS condition or time point.

For the tACS by time interaction, pairwise comparisons revealed that HD participants’ CNV amplitude significantly *increased* from pre-to-post alpha-tACS (pre-vs post-alpha mean difference = 0.80, *p = 0*.*01*). Conversely, the HD group’s CNV amplitude significantly *decreased* from pre-to-post sham (pre-vs post-sham mean difference = -0.40, *p = 0*.*03*). The HD group’s CNV amplitude did not change from pre-to-post delta-tACS (pre-vs post-delta mean difference = 0.11, *p = 0*.*64*).

With regards to differences between the tACS conditions at baseline and post-stimulation, pairwise comparisons found no significant differences between the three tACS conditions at baseline (alpha vs sham mean difference = 0.18, *p = 1*.*00*; alpha vs delta mean difference = 0.25, p = 0.85; delta vs sham = 0.-0.07, *p = 0*.*19*). For post-stimulation, however, there was a significant difference between the alpha and sham conditions (alpha vs sham mean difference = -1.01, p = 0.008), but not between alpha and delta (alpha vs delta mean difference = -0.43, *p = 0*.*38*) or delta and sham conditions (delta vs sham mean difference = -0.57, *p = 0*.*08*).

Overall, these results indicate that HD participants’ CNV amplitude for all three cue types became larger in response to alpha-tACS, but smaller in response to sham, while delta-tACS appeared to result in maintenance of CNV amplitude over time. Refer to Figures 3 and 4A-C.

**Figure 3.**
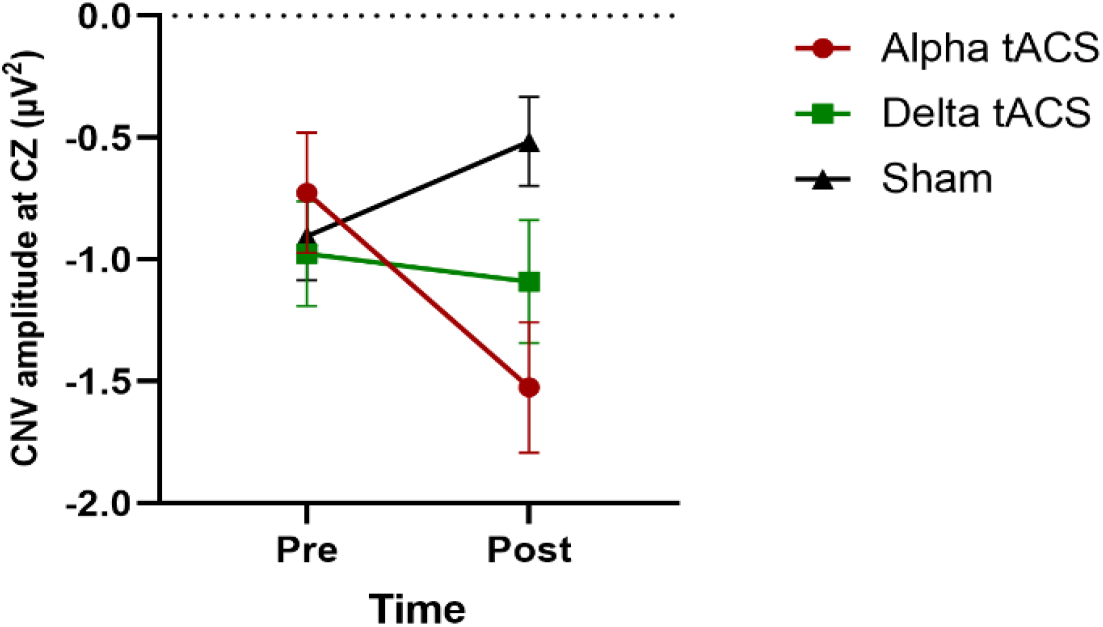
Graph of the significant interaction between tACS and time on CNV amplitude in the HD group (lines indicate standard error).

**Figure 4.**
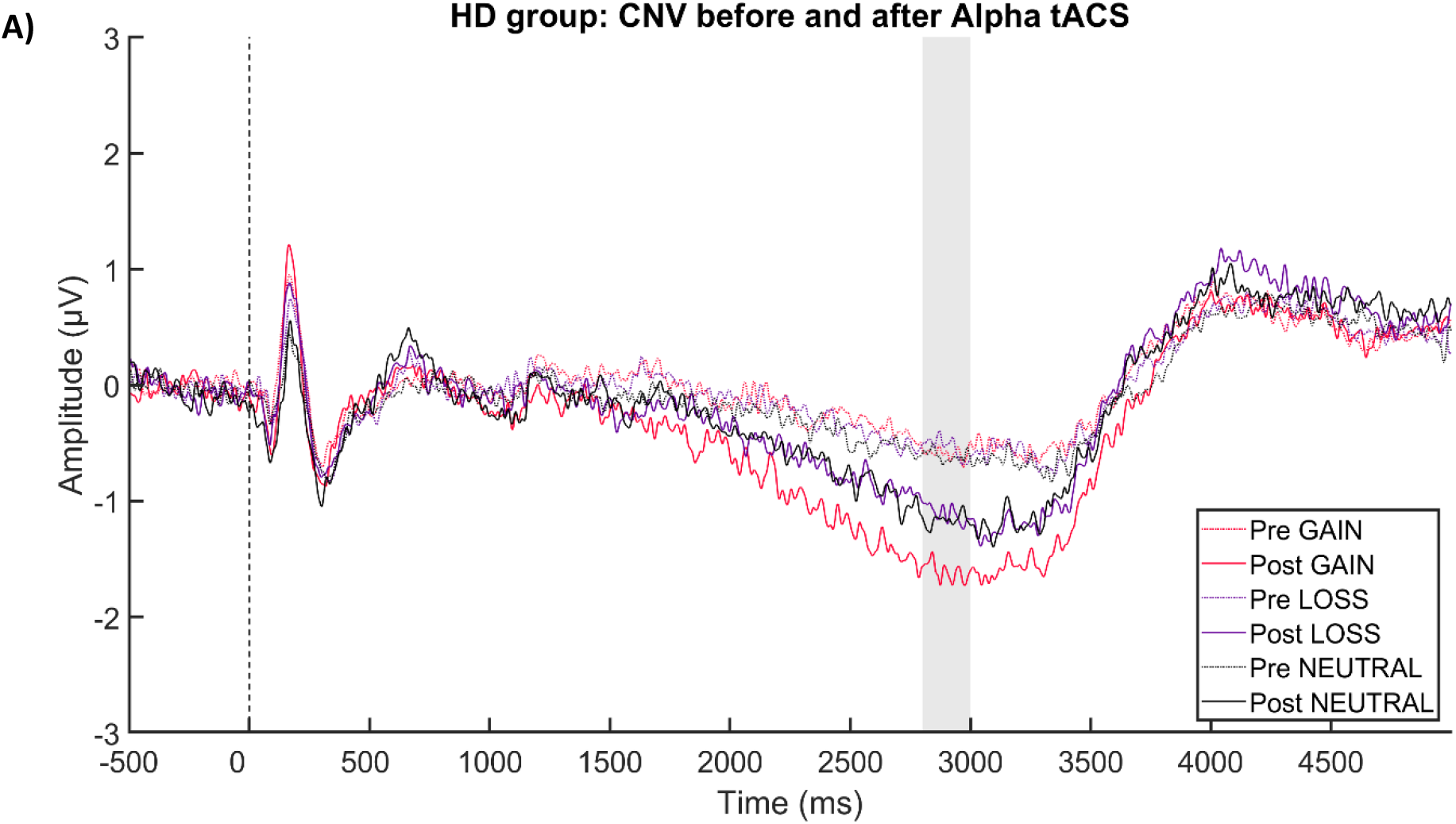
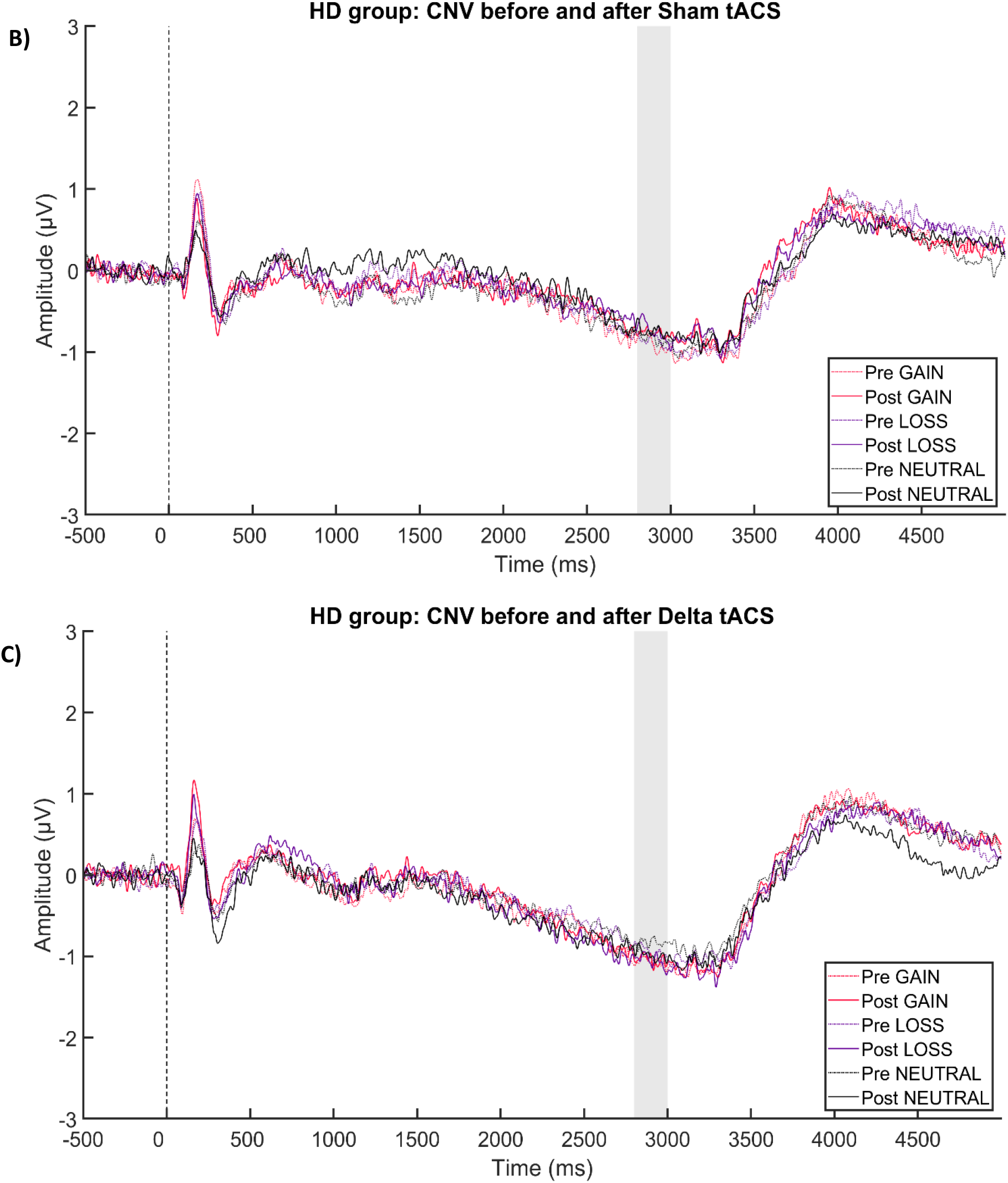
A) the ERP plot of HD participants’ CNV at electrode Cz for each cue type pre- (dotted lines) and post- (unbroken lines) alpha-tACS; B) pre- and post-sham-tACS; and C) pre- and post-delta-tACS. Grey bar indicates time window used for statistical analysis.

**Figure 5.**
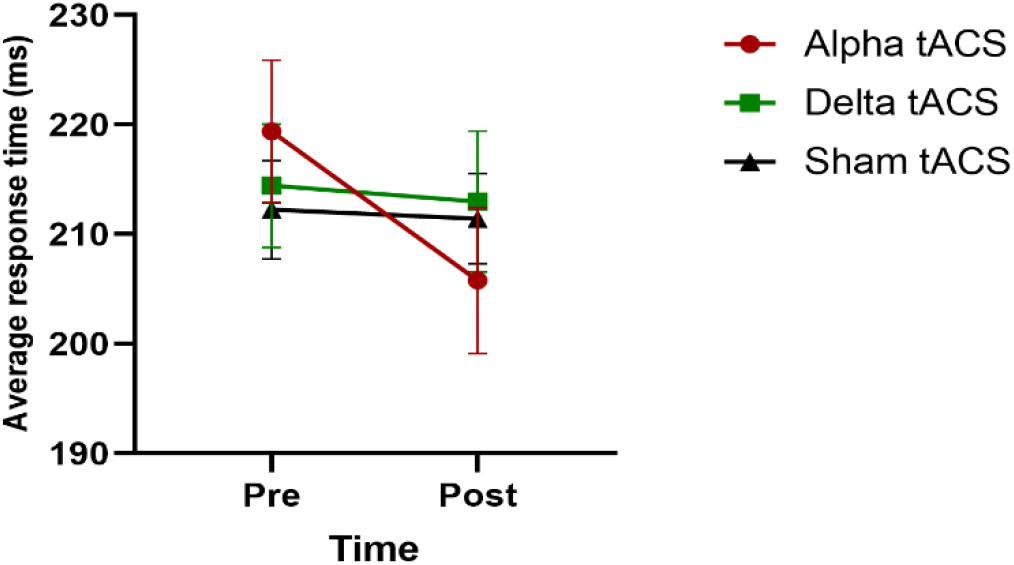
Graph of the significant interaction effect of tACS by time on average response times in the neurotypical control group (lines indicate standard error).

Given an IAF was not able to be detected for nearly half of the participants with HD, we conducted a mixed between-within subjects’ ANOVA just within the alpha-tACS condition (n = 21) to explore whether IAF detection affected HD participants’ CNV amplitude across the different cue types from pre-to-post alpha-tACS (*group:* IAF vs non-IAF *x cue type:* gain vs loss vs neutral x *time:* pre vs post). There was a significant main effect for time *(F1*,*19 = 8*.*55, p = 0*.*009, partial eta2 = 0*.*31*), confirming that CNV amplitude in response to all three cue types increased from pre-to-post alpha-tACS (mean difference = 0.72, *p = 009*). The between-subjects factor of IAF detection was, however, not significant *(F1*,*19 = 2*.*81, p = 0*.*11, partial eta2 = 0*.*13*), suggesting that receipt of tACS at the participant’s IAF versus at 10Hz did not significantly alter the modulatory effect of alpha-tACS on CNV amplitude. There were no other significant main or interaction effects.

Finally, as the CNV is sensitive to dopaminergic manipulation (Linssen et al. 2011), we also conducted a mixed between-within subjects ANOVA just within the alpha-tACS condition (n = 21) to explore the effect of risperidone use on participants’ CNV amplitude across the different cue types from pre-to-post alpha-tACS. With risperidone use (n = 7) as a between-subjects factor, there was still a significant main effect for time *(F1*,*19 = 9*.*50, p = 0*.*006, partial eta2 = 0*.*33*). The between-subjects factor of risperidone, however, was not significant *(F1*,*19 = 0*.*02, p = 0*.*89, partial eta2 = 0*.*001*). There were no other significant main or interaction effects. These results suggest that use of risperidone did not confound the effect of alpha-tACS on CNV amplitude.

#### 3.6.2 Neurotypical controls

Analysis of the effect of tACS on CNV amplitude in the control group revealed a significant main effect for cue type *(F2*,*18 = 8*.*24, p = 0*.*003, partial eta2 = 0*.*48*). There were no other significant main effects or interactions. Collapsed across tACS condition and time, pairwise comparisons showed that controls’ CNV amplitude following gain cues was significantly greater than that following neutral cues (gain vs neutral mean difference = -0.38, *p = 0*.*002*). Controls’ CNV amplitude in response to gain cues did not, however, differ significantly from their response to loss cues (gain vs loss mean difference = -0.14, *p = 0*.*33*), nor was there a significant difference in CNV amplitude elicited by loss cues relative to neutral cues (loss vs neutral mean difference = -0.24, *p = 0*.*06*). This indicates that controls’ CNV amplitude was greater in response to gain cues regardless of tACS condition or time point, and that neither delta-nor alpha-tACS modified the CNV in control participants.

### 3.7 Effect of tACS on P300 amplitude

The effect of tACS on P300 amplitude at electrode Pz following each cue type was assessed using separate within-subjects’ ANOVAs for each group (*tACS:* alpha vs delta vs sham x *cue type:* gain vs loss vs neutral x *time:* pre vs post).

#### 3.7.1 Participants with HD

There were no significant main effects or interactions for the HD group (n = 17). Again, we conducted a mixed between-within subjects ANOVA just within the alpha-tACS condition (n = 21) to explore whether IAF detection affected HD participants’ P300 amplitude across the different cue types from pre-to-post alpha-tACS (*group:* IAF vs non-IAF *x cue type:* gain vs loss vs neutral x *time:* pre vs post). The between-subjects factor of IAF detection was not significant *(F1*,*19 = 0*.*43, p = 0*.*52, partial eta2 = 0*.*02*), suggesting that receipt of tACS at the participant’s IAF versus 10Hz did not significantly alter the results for alpha-tACS on P300 amplitude.

#### 3.7.2 Neurotypical controls

The ANOVA for the control group revealed a significant main effect for cue type *(F2*,*18 = 10*.*30, p = 0*.*001, partial eta2* = 0.53). There were no other significant main effects or interactions. Collapsed across tACS condition and time, pairwise comparisons showed that controls’ P300 amplitude following gain and loss cues was significantly greater than that elicited by neutral cues (gain vs neutral mean difference = 0.60, *p = <0*.*001*; loss vs neutral mean difference = 0.41, *p = 0*.*003*). Controls’ P300 amplitude in response to gain cues, however, did not significantly differ from their response to loss cues (gain vs loss mean difference = 0.19, *p = 0*.*24*). This indicates that controls’ P300 amplitude was greater in response to motivationally salient cues, regardless of tACS condition or time point.

### 3.8 Effect of tACS on response times

The impact of tACS on average response times following each cue type was assessed using separate within-subjects’ ANOVAs for each group (*tACS:* alpha vs delta vs sham x *cue type:* gain vs loss vs neutral x *time:* pre vs post).

#### 3.8.1 Participants with HD

The ANOVA for the HD group (n = 18) revealed a significant main effect for time *(F1*,*17 = 7*.*83, p = 0*.*01, partial eta2 = 0*.*31*). Collapsed over tACS condition and cue type, pairwise comparisons found that HD participants’ average response time (in ms) significantly decreased from pre-to-post tACS across all tACS conditions (pre-vs post-tACS mean difference = 11.49, *p = 0*.*01*), suggesting within-session practice effects. There were no other significant main or interaction effects.

#### 3.8.2 Neurotypical controls

The ANOVA for the control group revealed a significant main effect for cue type *(F2*,*18 = 14*.*04, p = <0*.*001, partial eta2 = 0*.*61*), as well as a significant interaction for tACS by time *(F2*,*18 = 3*.*75, p = 0*.*04, partial eta2 = 0*.*29*). For the main effect of cue type, pairwise comparisons showed that controls’ average response times following gain cues and loss cues were both significantly faster than their response times following neutral cues (gain vs neutral mean difference = -8.30, *p = <0*.*001;* loss vs neutral mean difference = -7.11, *p = <0*.*001*). The comparison between response times following gain and loss cues was not significant (gain vs loss mean difference = - 1.16, *p = 0*.*17*). This indicates that controls responded more quickly following the presentation of motivationally salient cues, regardless of tACS condition or time point.

For the tACS by time interaction, pairwise comparisons revealed that controls’ average response time significantly *decreased* from pre-to-post alpha-tACS (pre-vs post-alpha mean difference = -13.60, *p = 0*.*002*). There was no significant change in controls’ average response time from pre-to-post delta-tACS (pre-vs post-delta mean difference = -1.45, p = *0*.*61*) or pre-to-post sham (pre-vs post-sham-tACS mean difference = -0.81, *p = 0*.*68*). Refer to Figure 4.

## 4.1 Discussion

In this proof-of-concept investigation of the effects of mPFC tACS for apathy in HD, HD participants’ CNV amplitude increased following tACS administered within the alpha frequency band, regardless of the motivational salience of the cue. Conversely, HD participants’ CNV amplitude across all three cue types decreased following sham-tACS but remained unchanged in response to tACS administered in the delta frequency band. Controls’ CNV amplitudes were significantly greater in response to motivationally salient cues, but this did not change as a function of tACS condition or time.

Contrary to expectations, neither the HD nor the control group demonstrated any change in P300 amplitude in any of the tACS conditions. The HD group’s average response times decreased across all cue types in all three tACS conditions, suggesting practice effects. By contrast, the neurotypical control group demonstrated significantly faster average response times following all cue types after receiving tACS administered within the alpha frequency band.

Blinding was effective and the stimulation was well-tolerated by both groups, with participants experiencing only minimal and transient side effects, consistent with previous tACS research (Matsumoto and Ugawa 2017).

### 4.2 CNV and P300 amplitudes

We significantly increased HD participants’ CNV amplitude using alpha-tACS. The association between the CNV and frontal systems, particularly nuclei within the CBGT networks (Plichta et al. 2013, Linssen et al. 2011), suggests successful engagement of our mPFC target. The converse effect of the sham condition, whereby HD participants’ CNV amplitude significantly *decreased* from pre-to-post sham suggests attenuation of the CNV secondary to habituation to the task and potential cognitive fatigue from the prolonged EEG session (Boksem, Meijman, and Lorist 2006). The non-significant effect of the delta-tACS condition aligns with this interpretation, in that participants still received 20 minutes of 2mA stimulation, albeit at a lower frequency, which may have *prevented* the attenuation of CNV amplitude that occurred over time in the sham condition.

The effect of alpha-tACS on HD participants’ CNV did not differentially modulate cues according to motivational salience. Such an interaction effect, whereby amplitude in response to gain and loss cues was enhanced relative to neutral cues, would have confirmed the specificity and relevance of our choice of neuroanatomical and oscillatory targets to motivation. Such specificity may, however, be unrealistic given the limited spatial precision of transcranial electrical stimulation techniques relative to other forms of brain stimulation (Polanía, Nitsche, and Ruff 2018).

We did not find a significant effect for IAF detection on the alpha tACS results in our HD group. Visual inspection of the CNV ERP plots suggest a differential response in participants for whom an IAF was detected (refer to Figures s1 and s2), but this could also reflect the difference in average DBS between the IAF subgroups; those participants for whom an IAF was detected having a lower DBS. Given the evidence supporting the use of individualised frequencies to maximise stimulation effects in neurotypical controls (Negahbani et al. 2018, Vogeti, Boetzel, and Herrmann 2022, Kemmerer et al. 2022), future research should attempt to understand the interaction between IAF detection and DBS to further refine stimulation protocols. There is also literature emerging on the use of ‘closed-loop’ tACS protocols, whereby the stimulation frequency updates in ‘real time’ based on features of the participant’s EEG to individualise input at all stages of stimulation (Frohlich and Townsend 2021). This ‘closed-loop’ approach may produce enhanced stimulation effects, but such personalised protocols for HD participants will not be possible without reliable individualised frequency detection.

Consistent with previous research, neurotypical control participants demonstrated larger CNV amplitudes in response to motivationally salient cues (Hill et al. 2018, Novak and Foti 2015, Pfabigan et al. 2014, Zhang et al. 2017), but this did not change as a function of tACS condition or time. This may reflect potential ‘ceiling effects’ on ERP amplitude in neurotypical controls or inadequate statistical power. Given that our intention was to *restore* HD participants’ neural responses to motivationally salient cues, our failure to significantly *enhance* the same neural responses in neurotypical controls is of scientific interest but does not detract from the clinical relevance of our findings for the HD group.

The failure to modulate the P300 amplitude in both groups may reflect the broad functional associations of this ERP, and its parietal (as well as frontal) spatial distribution (Luck 2014, Polich 2012), potentially making it less susceptible to tACS targeting motivational processes via mPFC. Nevertheless, given the anticipated effect of tACS on broader cortical oscillatory activity, particularly in response to alpha-tACS (Schutter and Wischnewski 2016, Klink et al. 2020), the lack of change in P300 amplitude was unexpected. Limited statistical power in combination with increased variability due to the broader age range of both groups may partly explain our negative findings; increased age having been found to correlate with lower ERP amplitude on the EEG version of the MID (Hill et al. 2018).

### 4.3 Response times

HD participants’ response times decreased from pre-to-post stimulation across all tACS conditions, suggesting within-session practice effects. This result was unexpected but may reflect HD participants’ difficulties with automatization of behaviours due to the involvement of the basal ganglia in procedural learning and automaticity (Thompson et al. 2010, Ashby, Turner, and Horvitz 2010). As such, many of our HD participants may, to some extent, have been re-learning the task (pressing the enter key in response to target presentation) each session, with incremental improvements in task execution as the session progressed, presenting as within-sessions practice effects. The latter may have also masked modulation of response times by tACS. By contrast, controls demonstrated the expected variability in response times as a function of cue motivational salience and improved their speed in response to all cue types from pre-to-post alpha-tACS (Hill et al. 2018, Novak and Foti 2015, Pfabigan et al. 2014, Zhang et al. 2017).

The incongruence between the ERP and response time results for both groups was unexpected given that the subjective valuation processes that precede behavioural output (i.e., registering the motivational salience of the cue and deciding how much effort to expend in executing a response) would be expected to influence the behavioural response (Chong 2018, Husain and Roiser 2018, Le Heron et al. 2018, Zhang and Zheng 2022). Indeed, using the EEG version of the MID in with neurotypical controls, Zhang and colleagues (2017) found that CNV amplitude was significantly positively correlated with response times across all three cue types, however, their sample size was significantly larger and more homogeneous than ours (i.e., 56 undergraduates aged between 17 and 23 years) (Zhang et al. 2017).

### 4.4 Limitations

These findings require replication with a larger sample to increase statistical power and enable refinement of the protocol for any clinical trials, particularly with regard to IAF detection and participant characteristics. Future studies also need to address the technological limitations that required the researcher to calculate and input participants’ IAF into the stimulation device, resulting in the current study being single rather than double blind. Inclusion of relevant behavioural measures before and after tACS administration for correlation with neurophysiological metrics, such as physical and cognitive effort discounting tasks (Atkins et al. 2020), would also increase confidence in the protocol’s therapeutic potential.

### 4.5 Conclusion

Apathy is a highly prevalent and burdensome symptom in HD, with evidence indicating irregular oscillatory activity and medial prefrontal and anterior cingulate cortices as pathophysiological targets for intervention. In this proof-of-concept study, we successfully increased HD participants’ CNV amplitude elicited during the EEG version of the MID task using alpha frequency tACS targeting the mPFC. We consider this preliminary evidence of tACS successfully modulating the pathophysiological circuits potentially underlying apathy in HD.

## Data Availability

De-identified data produced in the present study are available to genuine researchers upon reasonable request, provided confidentiality is maintained.

## Acknowledgements

MCD was supported by the Research Training Program Stipend and a Monash Graduate Excellence Scholarship. KEH was supported by National Health and Medical Research Council (NHMRC) Fellowships (1082894 and 1135558). PBF was supported by an Investigator Fellowship from the NHMRC (1193596). ATH was supported by an Alfred Deakin Postdoctoral Research Fellowship. We gratefully acknowledge the time, involvement, and feedback from the participants. This research would not have been possible without them. We also acknowledge the assistance with participant recruitment provided by the Statewide Progressive Neurological Disease Service at Calvary Health Care Bethlehem.

## Supplementary materials

**Table s1.** Comparison of baseline measures in HD participants according to diagnostic status.

|  |  |  |  |  |
| --- | --- | --- | --- | --- |
| Variable | Late premanifest | Early manifest | Statistics |  |
| Age in years (mean ±SD)<br>Range | 43.64 (±10.61)<br>23-63 | 53.55 (±10.04)<br>30-65 | $t_{20} = -2.25$ | $p = .04$ |
| Education years (mean ±SD)<br>Range | 14.36 (±2.38)<br>11-18 | 13.91 (±2.60)<br>11-20 | $t_{20} = 0.43$ | $p = .67$ |
| MoCA (median ±IQR)<br>Range | 26 (±4)<br>21-29 | 25 (±4)<br>19-29 | $U_{19} = 30.50$ | $p = .24$ |
| SDMT (mean ±SD)<br>Range | 47.45 (±13.63)<br>29-71 | 34.40 (±10.75)<br>14-53 | $t_{19} = 2.42$ | $p = .03$ |
| AES-C (median ±IQR)<br>Range | 27 (±5)<br>25-41 | 29 (±9)<br>24-53 | $U_{21} = 64.00$ | $p = .56$ |
| HADS – Anxiety (median ±IQR)<br>Range | 5 (±3)<br>1-8 | 2.50 (±6)<br>0-9 | $U_{22} = 51.50$ | $p = .56$ |
| HADS – Depression (median ±IQR)<br>Range | 1 (±2)<br>1-7 | 3 (±6)<br>0-8 | $U_{22} = 76.50$ | $p = .30$ |
| UHDRS-TMS (median ±IQR)<br>Range | 4 (±4)<br>2-16 | 14 (±27)<br>7-49 | $U_{19} = 81.50$ | $p = .001$ |
| DBS (mean ±SD)<br>Range | 351.05 (±60.32)<br>286-484.50 | 370.70 (±59.30)<br>283.50-495 | $t_{20} = 0.77$ | $p = .45$ |
| TFC (median ±IQR)<br>Range | 13 (±0)<br>12-13 | 10 (±3)<br>9-13 | $U_{22} = 18.50$ | $p = .004$ |
| Medication all types | 5 | 9 | $\chi^2 = 3.14$ | $p = .08$ |
| <sup>a</sup>Independent samples t-test – 2-tailed; <sup>b</sup>Chi-Square test; <sup>c</sup>Wilcoxon rank sum/Mann Whitney U test; SD = standard deviation; IQR = inter-quartile range; MoCA = Montreal Cognitive Assessment (max score = 30); SDMT = written version of the Symbol Digit Modalities Test – written version (max score = 110); AES-C = Apathy Evaluation Scale – Clinician version (scores are between = 18 - 72); HADS = Hospital Anxiety and Depression Scale (subscale max scores = 21); DBS = Disease Burden Score (CAG-35.5 x age); UHDRS – TMS = United Huntington's Disease Rating Scale® - Total Motor Scale (max score = 131); TFC = Total Functional Capacity score (scores are between 0 – 13) |  |  |  |  |

### IAF detection

Peak IAF was calculated in MATLAB using a customized script. Five-minute resting state EEG recordings were segmented into four second epochs overlapping 25%, before being preprocessed and cleaned in a fully automated fashion utilizing the Harvard Automated Processing Pipeline for Electroencephalography (HAPPE) (Gabard-Durnam et al. 2018).

To reduce overall processing time the HAPPE pipeline was modified to use the fastICA algorithm (Hyvarinen 1999) in place of the default ICA algorithm used by HAPPE for the decomposition of independent components.

Channel Fz was used to estimate power in alpha frequency band (8-12Hz) for each epoch based on a power spectral density estimate (pwelch method). To ensure reliable alpha peak detection we automatically discarded 15% of epochs that returned lowest alpha power values.

Individual alpha frequency (IAF) estimation was then calculated using the remaining high alpha power epochs, and a selection of frontocentral electrodes (F7, F5, F3, F1, Fz, F2, F4, F6, F8, FC3, FC1, FC2, FC4), this method of reliable alpha peak detection employs Savitzky-Golay filter (SGF) as described by Corcoran and colleagues (2018).

**Table s2.**
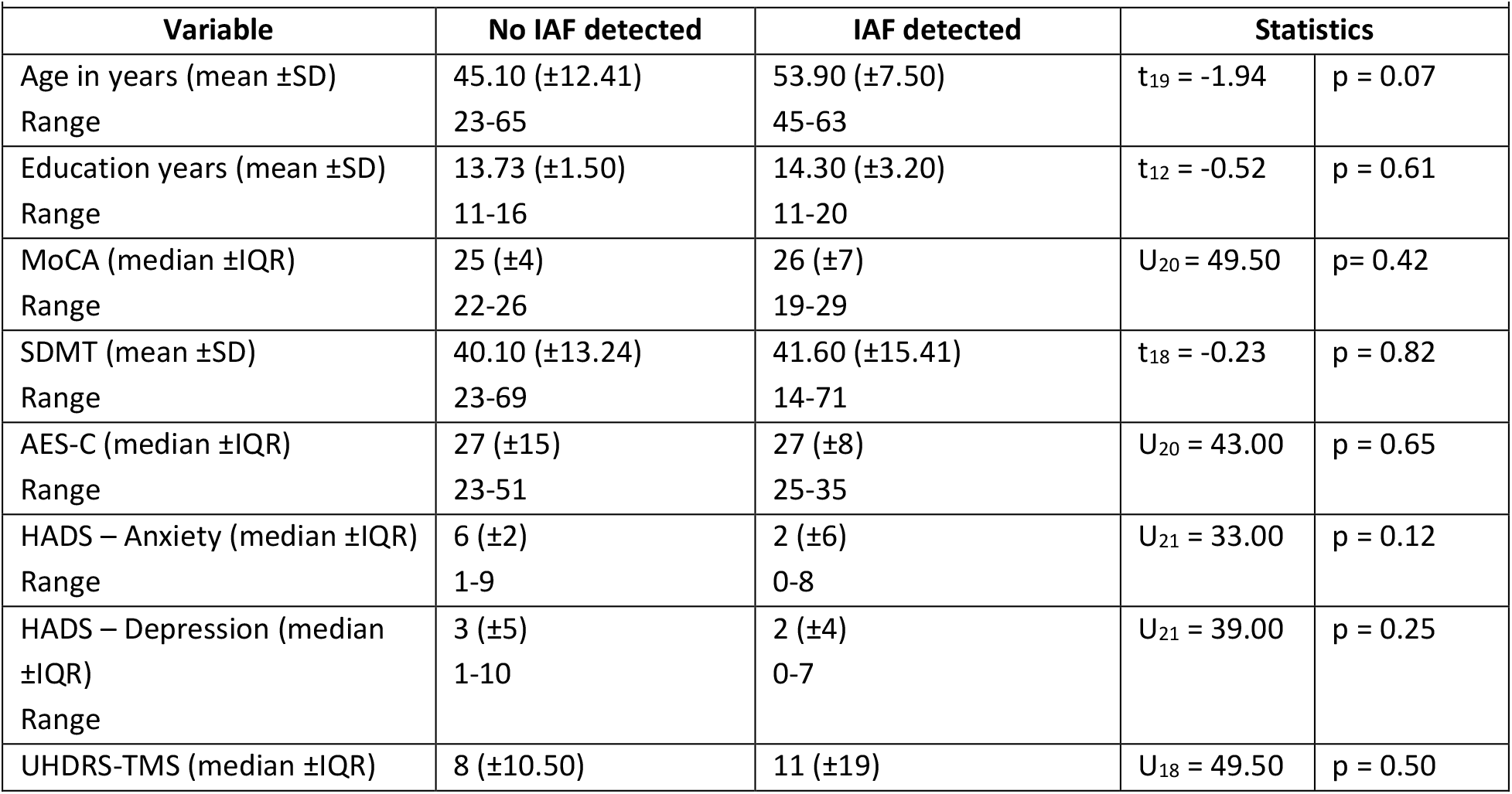
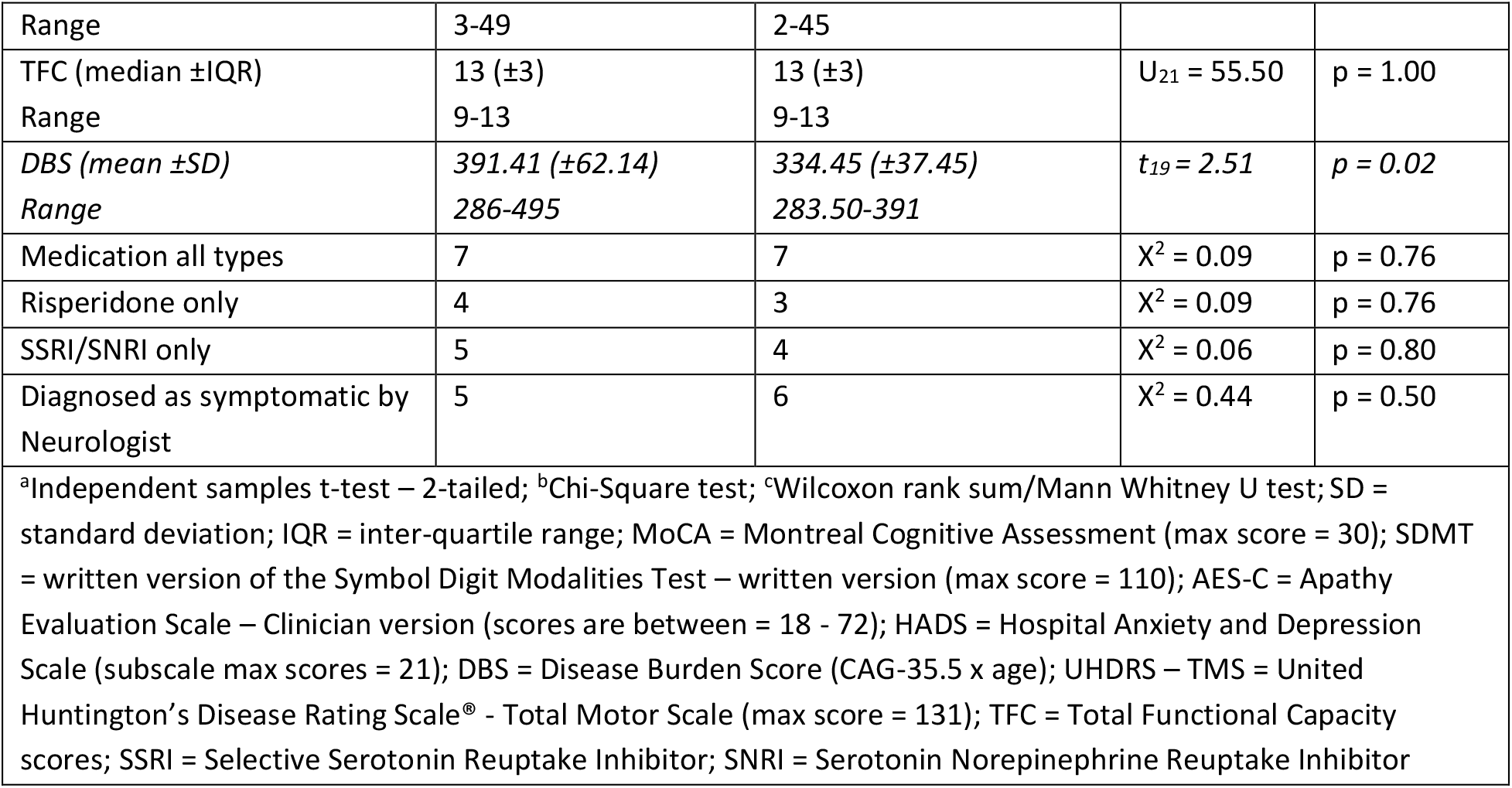
Comparison of baseline measures in HD participants according to IAF detection.

|  |  |  |  |  |
| --- | --- | --- | --- | --- |
| **Variable** | **No IAF detected** | **IAF detected** | **Statistics** |  |
| Age in years (mean ±SD)<br>Range | 45.10 (±12.41)<br>23-65 | 53.90 (±7.50)<br>45-63 | $t_{19} = -1.94$ | $p = 0.07$ |
| Education years (mean ±SD)<br>Range | 13.73 (±1.50)<br>11-16 | 14.30 (±3.20)<br>11-20 | $t_{12} = -0.52$ | $p = 0.61$ |
| MoCA (median ±IQR)<br>Range | 25 (±4)<br>22-26 | 26 (±7)<br>19-29 | $U_{20} = 49.50$ | $p = 0.42$ |
| SDMT (mean ±SD)<br>Range | 40.10 (±13.24)<br>23-69 | 41.60 (±15.41)<br>14-71 | $t_{18} = -0.23$ | $p = 0.82$ |
| AES-C (median ±IQR)<br>Range | 27 (±15)<br>23-51 | 27 (±8)<br>25-35 | $U_{20} = 43.00$ | $p = 0.65$ |
| HADS – Anxiety (median ±IQR)<br>Range | 6 (±2)<br>1-9 | 2 (±6)<br>0-8 | $U_{21} = 33.00$ | $p = 0.12$ |
| HADS – Depression (median ±IQR)<br>Range | 3 (±5)<br>1-10 | 2 (±4)<br>0-7 | $U_{21} = 39.00$ | $p = 0.25$ |
| UHDRS-TMS (median ±IQR) | 8 (±10.50) | 11 (±19) | $U_{18} = 49.50$ | $p = 0.50$ |
| Range | 3-49 | 2-45 |  |  |
| TFC (median ±IQR) | 13 (±3) | 13 (±3) | $U_{21} = 55.50$ | $p = 1.00$ |
| Range | 9-13 | 9-13 |  |  |
| DBS (mean ±SD) | 391.41 (±62.14) | 334.45 (±37.45) | $t_{19} = 2.51$ | $p = 0.02$ |
| Range | 286-495 | 283.50-391 |  |  |
| Medication all types | 7 | 7 | $X^2 = 0.09$ | $p = 0.76$ |
| Risperidone only | 4 | 3 | $X^2 = 0.09$ | $p = 0.76$ |
| SSRI/SNRI only | 5 | 4 | $X^2 = 0.06$ | $p = 0.80$ |
| Diagnosed as symptomatic by Neurologist | 5 | 6 | $X^2 = 0.44$ | $p = 0.50$ |

<sup>a</sup>Independent samples t-test – 2-tailed; <sup>b</sup>Chi-Square test; <sup>c</sup>Wilcoxon rank sum/Mann Whitney U test; SD = standard deviation; IQR = inter-quartile range; MoCA = Montreal Cognitive Assessment (max score = 30); SDMT = written version of the Symbol Digit Modalities Test – written version (max score = 110); AES-C = Apathy Evaluation Scale – Clinician version (scores are between = 18 - 72); HADS = Hospital Anxiety and Depression Scale (subscale max scores = 21); DBS = Disease Burden Score (CAG-35.5 x age); UHDRS – TMS = United Huntington's Disease Rating Scale® - Total Motor Scale (max score = 131); TFC = Total Functional Capacity scores; SSRI = Selective Serotonin Reuptake Inhibitor; SNRI = Serotonin Norepinephrine Reuptake Inhibitor

**Table s3.** Percentage of ‘correct’ responses in each condition before and after tACS according to group: Descriptive statistics for Winsorized data.

| Variable | Neurotypical controls | Participants with HD |
| --- | --- | --- |
| Pre-alpha tACS (mean ±SD) | 62 (±4.60) | 63 (±6.40) |
| Range | 57-80 | 56-85 |
| Post-alpha tACS (mean ±SD) | 61 (±1.80) | 61.70 (±3.97) |
| Range | 58-64 | 57-72 |
| Pre-delta tACS (mean ±SD) | 61.10 (±1.77) | 61.80 (±2.70) |
| Range | 59.66 | 58-67 |
| Post-delta tACS (mean ±SD) | 62.40 (±5.11) | 63.80 (±4.42) |
| Range | 58-83 | 59 - 74 |
| Pre-sham (mean ±SD) | 61.35 (±3.30) | 62.24 (±4.66) |
| Range | 58-73 | 57-81 |
| Post-sham (mean ±SD) | 61.30 (±2.51) | 63.10 (±5.32) |
| Range | 58-69 | 57-81 |

**Table s4.**
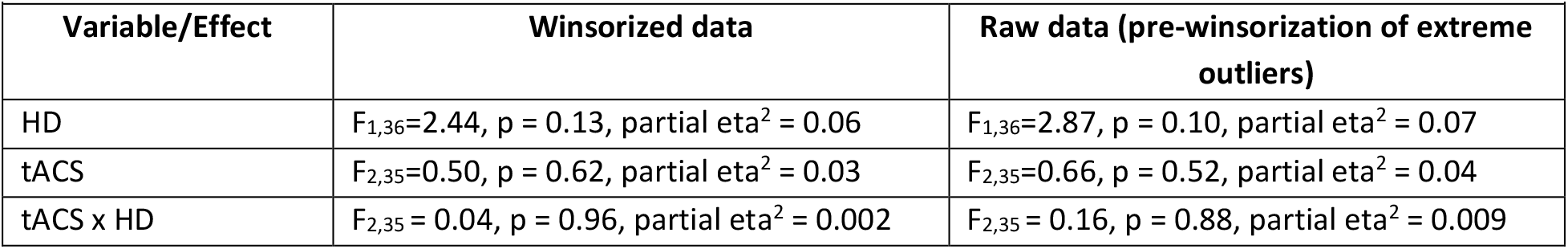
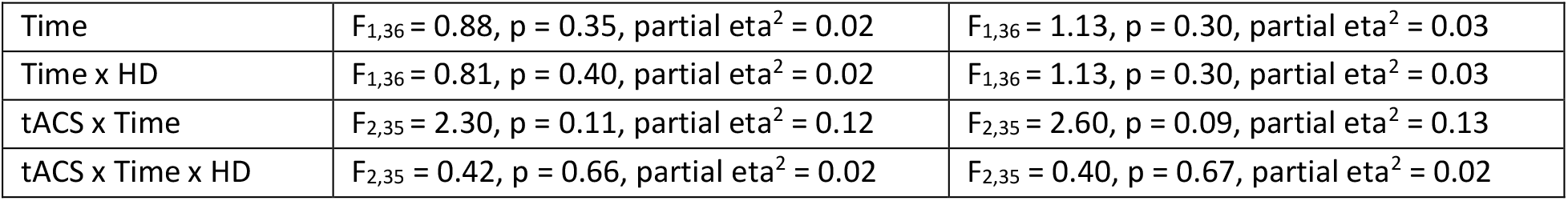
Percentage of ‘correct’ responses in each condition before and after tACS according to group: ANOVA output.

| Variable/Effect | Winsorized data | Raw data (pre-winsorization of extreme outliers) |
| --- | --- | --- |
| HD | $F_{1,36}=2.44$, $p = 0.13$, partial $\eta^2 = 0.06$ | $F_{1,36}=2.87$, $p = 0.10$, partial $\eta^2 = 0.07$ |
| tACS | $F_{2,35}=0.50$, $p = 0.62$, partial $\eta^2 = 0.03$ | $F_{2,35}=0.66$, $p = 0.52$, partial $\eta^2 = 0.04$ |
| tACS x HD | $F_{2,35} = 0.04$, $p = 0.96$, partial $\eta^2 = 0.002$ | $F_{2,35} = 0.16$, $p = 0.88$, partial $\eta^2 = 0.009$ |
| Time | $F_{1,36} = 0.88, p = 0.35, \text{partial } \eta^2 = 0.02$ | $F_{1,36} = 1.13, p = 0.30, \text{partial } \eta^2 = 0.03$ |
| Time x HD | $F_{1,36} = 0.81, p = 0.40, \text{partial } \eta^2 = 0.02$ | $F_{1,36} = 1.13, p = 0.30, \text{partial } \eta^2 = 0.03$ |
| tACS x Time | $F_{2,35} = 2.30, p = 0.11, \text{partial } \eta^2 = 0.12$ | $F_{2,35} = 2.60, p = 0.09, \text{partial } \eta^2 = 0.13$ |
| tACS x Time x HD | $F_{2,35} = 0.42, p = 0.66, \text{partial } \eta^2 = 0.02$ | $F_{2,35} = 0.40, p = 0.67, \text{partial } \eta^2 = 0.02$ |

**Figure s1.**
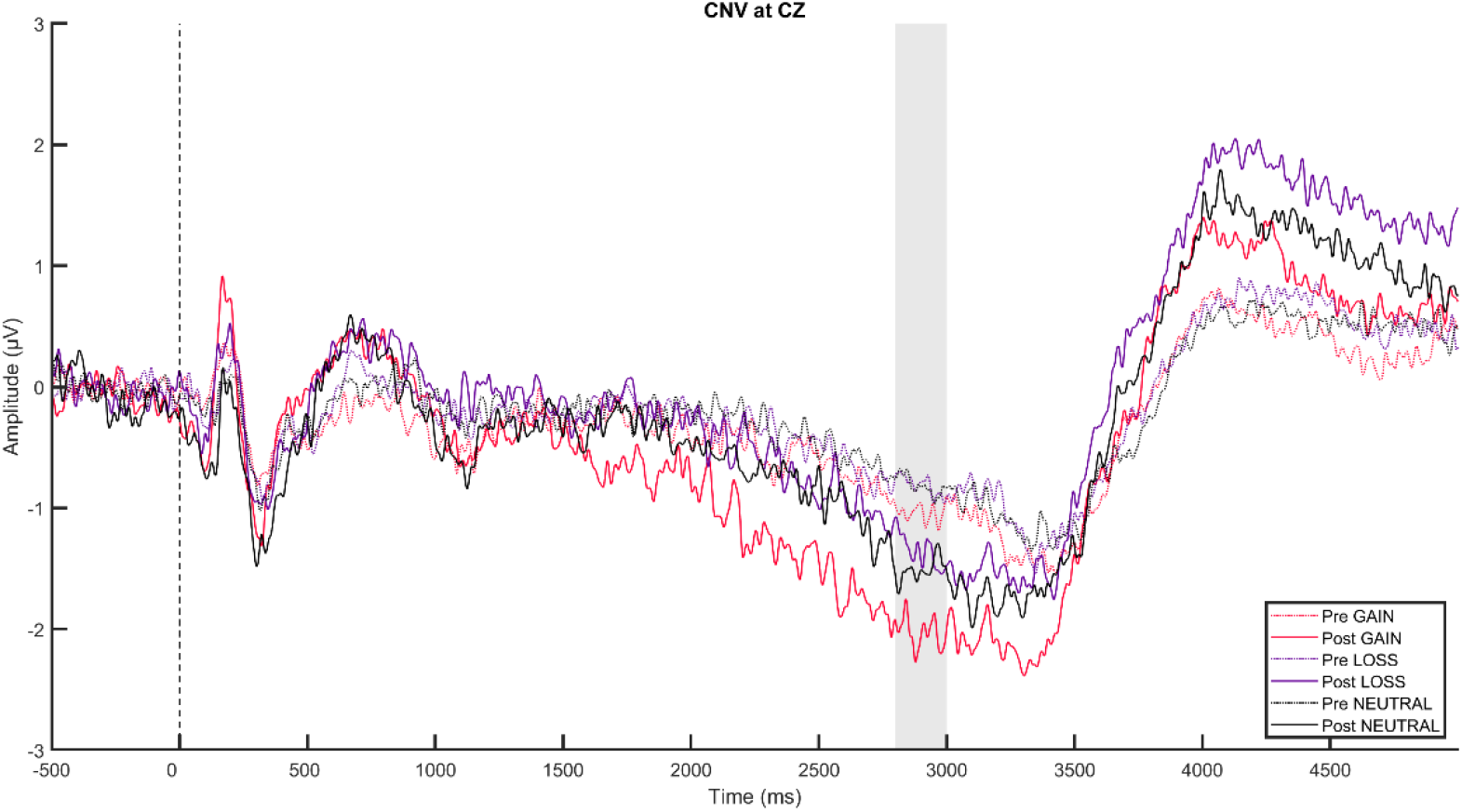
ERP plot of CNV pre and post alpha tACS of HD participants for whom an *IAF was detected*.

**Figure s2.**
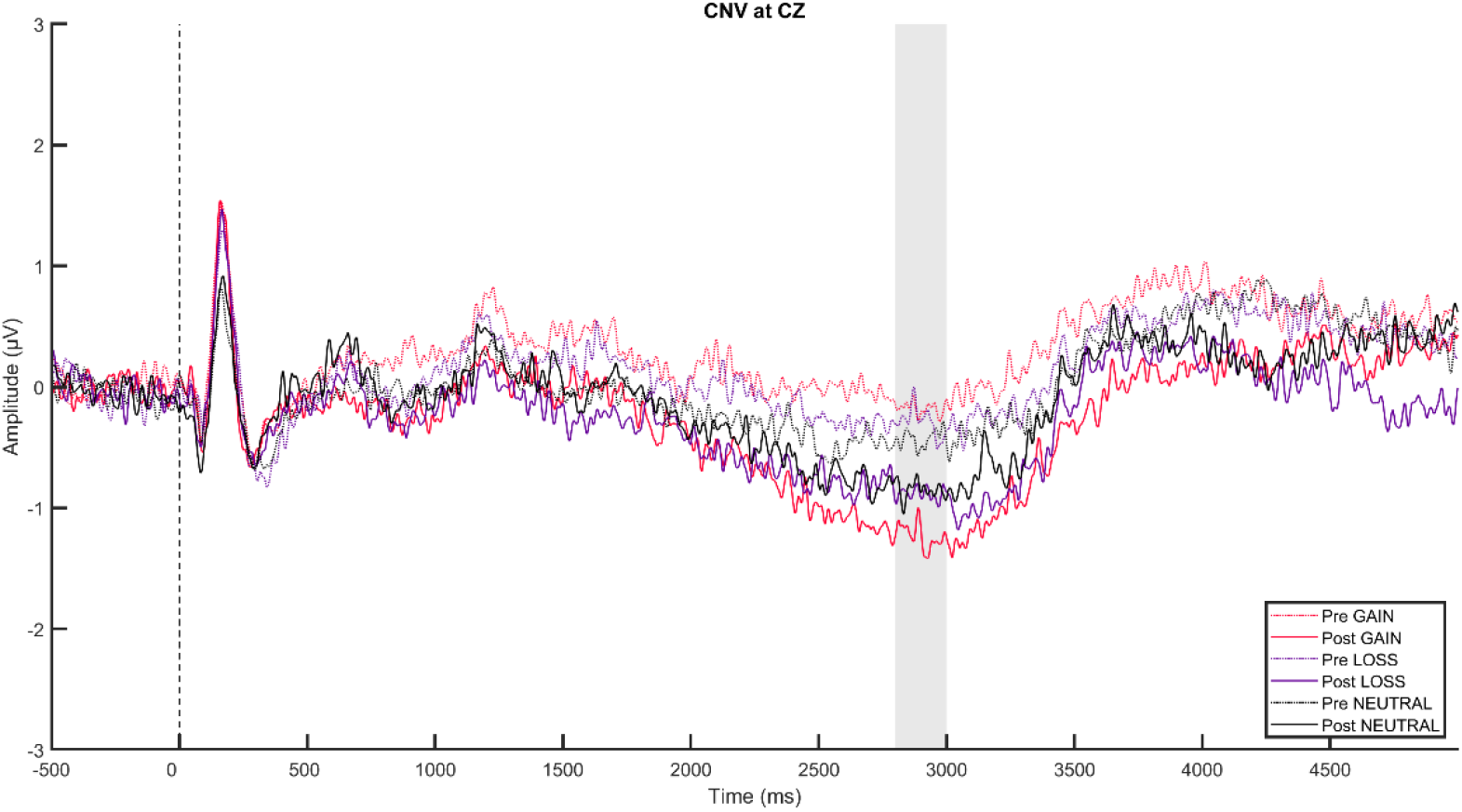
ERP plot of CNV pre and post alpha tACS of HD participants who received 10Hz (*IAF not detected)*

